# The impact of school reopening on the spread of COVID-19 in England

**DOI:** 10.1101/2020.06.04.20121434

**Authors:** Matt J. Keeling, Michael J. Tildesley, Benjamin D. Atkins, Bridget Penman, Emma Southall, Glen Guyver-Fletcher, Alex Holmes, Hector McKimm, Erin E. Gorsich, Edward M. Hill, Louise Dyson

**Affiliations:** The Zeeman Institute for Systems Biology & Infectious Disease Epidemiology Research, School of Life Sciences and Mathematics Institute, University of Warwick, Coventry, CV4 7AL, United Kingdom; Mathematics for Real World Systems Centre for Doctoral Training, Mathematics Institute, University of Warwick, Coventry, CV4 7AL, United Kingdom; Midlands Integrative Biosciences Training Partnership, School of Life Sciences, University of Warwick, Coventry, CV4 7AL, United Kingdom; Department of Statistics, University of Warwick, Coventry, CV4 7AL, United Kingdom

## Abstract

By mid-May, cases of COVID-19 in the UK had been declining for over a month; a multi-phase emergence from lockdown was planned, including a scheduled partial reopening of schools on 1st June. Although evidence suggests that children generally display mild symptoms, the size of the school-age population means the total impact of reopening schools is unclear. Here, we present work from mid-May that focused on the imminent opening of schools and consider what these results imply for future policy.

We compared eight strategies for reopening primary and secondary schools in England. Modifying a transmission model fitted to UK SARS-CoV-2 data, we assessed how reopening schools affects contact patterns, anticipated secondary infections and the relative change in the reproduction number, R. We determined the associated public health impact and its sensitivity to changes in social-distancing within the wider community.

We predicted reopening schools with half-sized classes or focused on younger children was unlikely to push *R* above one. Older children generally have more social contacts, so reopening secondary schools results in more cases than reopening primary schools, while reopening both could have pushed *R* above one in some regions. Reductions in community social-distancing were found to outweigh and exacerbate any impacts of reopening. In particular, opening schools when the reproduction number *R* is already above one generates the largest increase in cases.

Our work indicates that while any school reopening will result in increased mixing and infection amongst children and the wider population, reopening schools alone in June was unlikely to push *R* above one. Ultimately, reopening decisions are a difficult trade-off between epidemiological consequences and the emotional, educational and developmental needs of children. Into the future, there are difficult questions about what controls can be instigated such that schools can remain open if cases increase.

## Introduction

The emergence of a novel strain of coronavirus, now named SARS-CoV-2, in Wuhan city, China, in late 2019, has resulted in a global pandemic that spread to every region in the world. When the SARS-CoV-2 virus infects humans it can result in COVID-19 disease, with symptoms including a fever, a continuous dry cough, a shortness of breath and a loss of sense of taste and smell [1]. In severe cases, the symptoms can require hospitalisation and admission to intensive care, with ventilation required in the most severe cases in order to assist with breathing.

As the number of confirmed cases increased both nationally and globally, there was a concern that hospital and intensive care capacities would be rapidly overwhelmed without the introduction of interventions to curb the spread of infection. With this in mind, many countries introduced a range of social distancing measures, such as the closing of workplaces, pubs and restaurants, the restriction of leisure activities and the closing of schools. In the UK, the introduction of many of these measures was announced during the week of 16th March, with schools, along with the hospitality sector, closing on Friday 20th March. Full lockdown measures were subsequently introduced three days later, on the evening of Monday 23rd March. When we completed this work in late May, over 270,000 people in the UK had been confirmed to have been infected with COVID-19, with over 37, 500 confirmed deaths of individuals who had tested positive for infection.

The decision to close schools is a balance between the risk associated with transmission in the school environment and the educational and welfare impact upon children of shutting down education establishments. Evidence from a range of sources suggests that children are, in general, only mildly affected by the disease and have low mortality rates [2, 3]. This is reflected in the fact that by 27th May 2020 there had been 26,235 COVID-19 associated deaths in hospitals in England, but only 16 of those were in the 0-19 year age group [4]. In a retrospective study of 2,135 paediatric COVID-19 cases in China [5], 89.7% of children had mild or moderate disease while 5.8% were severe or critical; similarly low levels of severe disease are reported in other regions [3, 6]. The health risks of school attendance for any individual child is therefore thought to be low.

However, there is less certainty regarding children’s role in the transmission of SARS-CoV-2 [7, 8]. This can be broken down into two key questions: (i) how likely are children to become infected, and (ii) once infected, are children likely to transmit infection?

A meta-analysis concluded that children and young people under the age of 20 may be less likely to become infected: the odds ratio for becoming infected upon contact with an index case compared to adults (> 20 years old) is 0.44 (CI 0.29,0.69) [7]. This conclusion is based on pooling the results of contact tracing and population-screening studies, most of which find evidence that the attack rate in children may be lower than in adults [9, 10], but one does not (Bi *et al*. [11]). All contact tracing studies are hampered by the problem that symptom-based surveillance is likely to systematically under detect cases in children [11]. Seroprevalence surveys so far do not find any significant effect of age on the probability of possessing antibodies against COVID-19, although those under the age of five are not always included in surveys [12-14]. Two cross sectional PCR studies hint at lower susceptibility in children, since they found no SARS-CoV-2 PCR positive children under the age of 10 [15, 16], but a PCR-based survey by the UK Office for National Statistics found no difference in the probability of infection between age classes [17]. Further, large-scale seroprevalence studies which fully sample all age groups will be necessary to fully resolve these questions. Overall the balance of evidence cautiously suggests that children may have a lower inherent susceptibility. If it exists, such lower susceptibility could be physiological [18] or could be due to cross reactive immune responses from other childhood infections, with cross-protection between other human coronaviruses and SARS-CoV-2 hinted at by recent studies [19, 20].

There is little evidence from contact tracing and clinical investigations about the relative infectiousness of children. Children hospitalised with COVID-19 readily shed the virus above the likely transmission threshold [21-23], with detection of virus in nasopharyngeal (nasal) swabs, oropharyngeal (throat) swabs, sputum, or faeces [24, 25]. However, in their review of contact tracing and population-screening studies, Viner *et al*. [7] found just one relevant study comparing infectiousness by age: Zhu *et al*. [26], which shows that children make up a low proportion of index cases in households. As pointed out by Viner *et al*., this particular result could be explained by children being less likely to get infected in the first place rather than children being less infectious once they have actually contracted the virus. There is also evidence suggesting that mild cases in adults could be less infectious than severe or critical cases [10], but it remains unknown whether this result extends to asymptomatic or mild cases in children. Thus, children with severe symptoms are likely infectious, but it is harder to determine how transmissible the virus may be from children with few or no symptoms.

As of May 2020, we were aware of three reported studies of SARS-CoV-2 infection within the school environment. A retrospective serology study of 661 individuals with links to a school-based outbreak in Oise, France, showed that the infection spread readily within and outside the school to reach students, teachers, staff, and families [27]. In contrast, an Australian government study of cases in schools in Western Australia [28] identified nine children and nine adults who tested positive for SARS-CoV-2 (located across different schools), but found only two secondary cases when testing a third of the close contacts of these cases (288 samples). In Ireland, six SARS-CoV-2 cases were identified who had attended or taught in schools. None of 924 school related child contacts or 101 school related adult contacts showed any symptoms, but asymptomatic cases could have been missed [29]. The Australian school cases were identified between 5th March and 3rd April, and the first Irish school case was identified at the beginning of March. The first Oise school cases, by contrast, were identified on the 2nd February 2020. The greater awareness of COVID-19 by March, during which the WHO declared COVID-19 as a global pandemic, likely helped to control the Australian and Irish school-based outbreaks sooner than in Oise.

In the UK, during late May 2020, cases of COVID-19 were declining and there was strong evidence to suggest that the effective reproduction number (*R*) had dropped below 1 across the country. A multiphase relaxation plan for the country to emerge from lockdown began on 13th May, with a greater emphasis on returning to work if practical. We present here research formulated to address policy questions in May, to help inform the expected impact of various groups returning to the classroom. In particular, we investigate the epidemiological impacts of reopening schools in England, focusing on different combinations of year groups. We extend a previously developed dynamic transmission model for SARS-CoV-2, which is fit (on a regional basis for the UK) to real-time data on confirmed cases requiring hospital care and mortality. We compare and contrast multiple possible strategies for reopening both primary and secondary schools, focusing upon determining the effect of given year groups returning to school upon future epidemic behaviour. By elucidating the risks associated with particular age groups returning to school, we seek to contribute to the evidence base on the likely role of schools in the containment and control of this outbreak. Unlike other modelling studies [30], we decouple school reopening from other measures (such as a greater return to work); we feel this generates a clearer picture of the roles of school children and adults [31].

In England, primary schools partially reopened on 1st June: reception, year 1 and year 6 children initially returned, with an emphasis on maintaining social distancing measures where possible. In September (August in Scotland), the majority of schools reopened with generally high levels of attendance. We therefore discuss the implications for this work both in terms of the likely effects of schools on the unfolding epidemic and their role in any future imposition of additional control measures.

## Methods

### Transmission model

In order to perform the analysis of school reopening, we extended a previously-developed deterministic, age-structured compartmental SARS-CoV-2 transmission model [32]. The model was matched to a variety of data sources including hospitalisations, ICU occupancy and deaths, while age-dependent parameters were scaled to achieve agreement with the early age-distributions [33]. We stratified the population according to current disease status, following a susceptible-exposed-infectious-recovered (SEIR) paradigm (Fig. 1). We assumed the latent period to be Erlang distributed, modelled within the compartmental framework via division of the latent state into three stages. Infectious cases were partitioned by presence of symptoms, meaning we tracked symptomatic and asymptomatic individuals separately. Additional layers of complexity included differentiating by isolation and household status. We provide a listing of model parameters in Table 1, with a description of the model equations given in Supporting Text S1. We use the predicted number of symptomatic individuals to estimate the number of hospital admissions, ICU admissions and deaths, by estimating the proportion of symptomatic individuals requiring hospitalisation, ICU admission and the proportion that eventually die, and the distribution of times through each of these states. For hospital admissions and cases requiring treatment in ICU, the proportions going through each state and the distribution of times taken were drawn from the COVID-19 Hospitalisation in England Surveillance System (CHESS) data set that collects detailed data on patients infected with COVID-19 [34]. The risk of death was also captured with an age-dependent probability, while the distribution of delays between hospital admission and death was assumed to be age-independent, with both these two quantities determined from the Public Health England (PHE) death records.

**Fig. 1:**
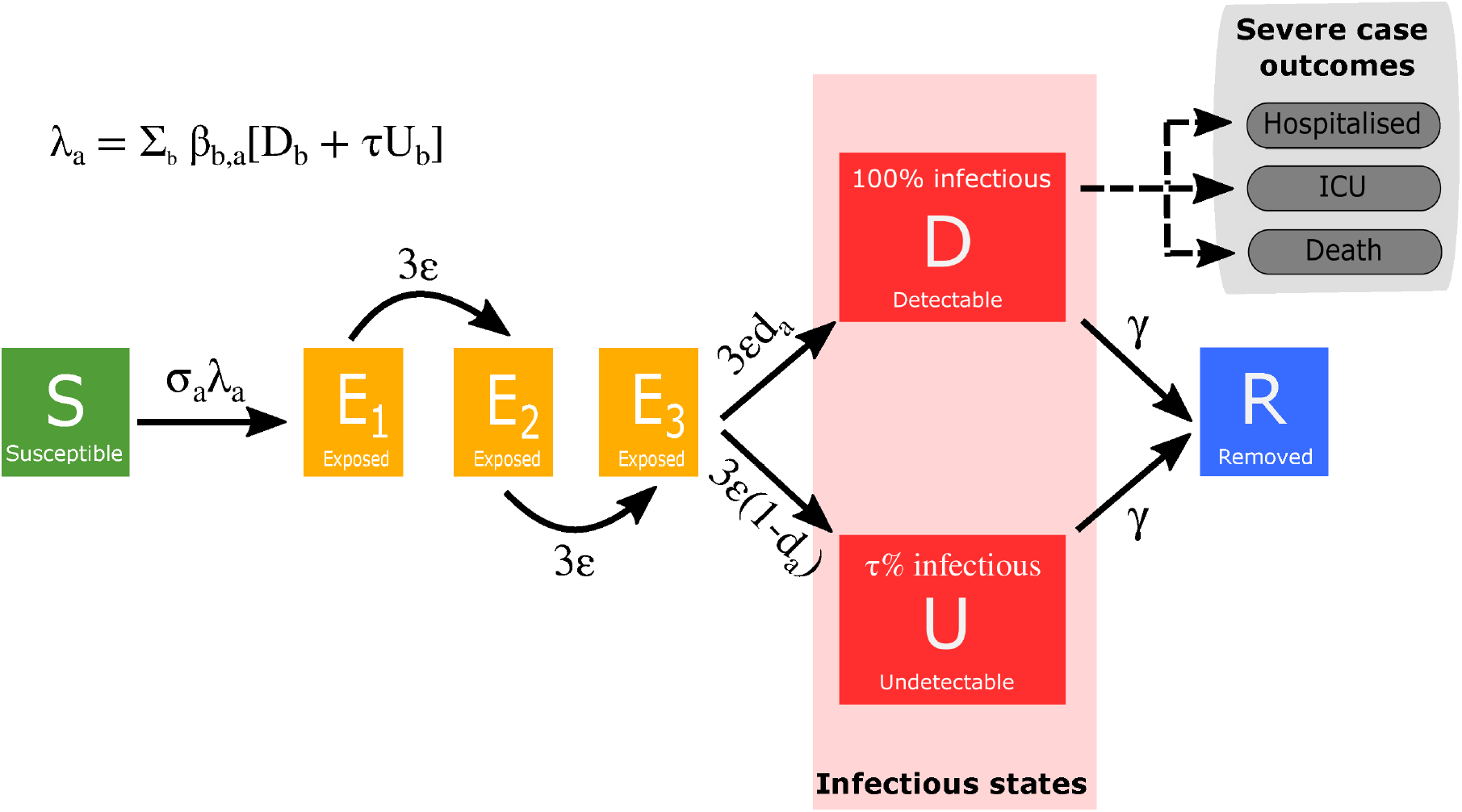
Disease states and transitions. We stratified the population into susceptible, exposed, detectable infectious, undetectable infectious, and removed states. Solid lines correspond to disease state transitions, with dashed lines representing mapping from detectable cases to severe clinical cases that require hospital treatment, critical care (ICU), or result in death. We separated those aged between 0 and 19 years old into single years, with the remainder of the population stratified into five year age brackets. See Table 1 for a listing of model parameters. Note, we have not included quarantining or household status on this depiction of the system.

**Table 1:** Key model parameters

| Parameter | Description | Value | Source |
| --- | --- | --- | --- |
| $\beta$ | Age-dependent transmission, split into household, school, work and other | | Derived from POLYMOD matrices [36] |
| $\epsilon$ | Rate of progression to infectious disease ($1/\epsilon$ is the duration in the exposed class) | $\sim 0.2$ | Fitted as part of MCMC process |
| $\gamma$ | Recovery rate, changes with $\tau$, the relative level of transmission from undetected asymptomatics compared to detected symptomatics | $\sim 0.5$ | Fitted from early age-stratified UK case data |
| $\alpha$ | Scales the degree to which age-structured heterogeneity is due to age-dependent probability of symptoms ($\alpha = 0$) or age-dependent susceptibility ($\alpha = 1$) | 0.137(0.1150.146) | Fitted as part of MCMC process |
| $\tau$ | Relative level of transmission from asymptomatic compared to symptomatic infection | 0.138(0.135 – 0.145) | Fitted as part of MCMC process |
| $d_a$ | Age-dependent probability of displaying symptoms (and hence being detected), changes with $\alpha$ and $\tau$ | 0-1 | Fitted from early age-stratified UK case data (see of MCMC process or varied according to scenario (see Supporting Figure S1) |
| $\sigma_a$ | Age-dependent susceptibility, changes with $\alpha$ and $\tau$ | 0.4-1 | Fitted from early age-stratified UK case data (see of MCMC process or varied according to scenario (see Supporting Figure S1) |
| $\phi^R$ | Adherence to the lockdown restrictions | 0.3 – 0.8 | Fitted as part of MCMC process or varied according to scenario (see Supporting Figure S1) |
| $H^R$ | Household quarantine proportion | 0 – 1 | Can be varied according to scenario |
| $N_a^R$ | Population size of a given age | By region | ONS |

With the inclusion of age-structure, transmission was governed through age-dependent mixing matrices, based on UK social mixing patterns [35, 36], scaled by an age-dependent susceptibility that was determined to produce the early age-distribution of symptomatic cases. To capture the effects of social distancing measures that were introduced in the UK to reduce transmission, we scaled down the mixing matrices associated with schools, work and other activities while increasing the within household transmission matrix (see Supporting Text S2).

In a refinement to the base model, we imposed an amended age-stratification of the population. Whilst in previous work the population was stratified into five year age brackets, for this study we separated those aged between 0 and 19 years old into single year cohorts, with the remainder of the population stratified into five year age brackets as before (20-24yrs, 25-29yrs and so on). The final age category corresponded to those aged 100 years or above. This fine-scale structure for those younger than 20 is important to be able to capture different policy questions; however resolution at a single year of age is not captured within the mixing matrices [35, 36]. We therefore generally retain the mixing structure based on five year age groups (Fig. 2), but assume that 70% of mixing within the same five year age group comes from interactions within the same school year.

**Fig. 2:**
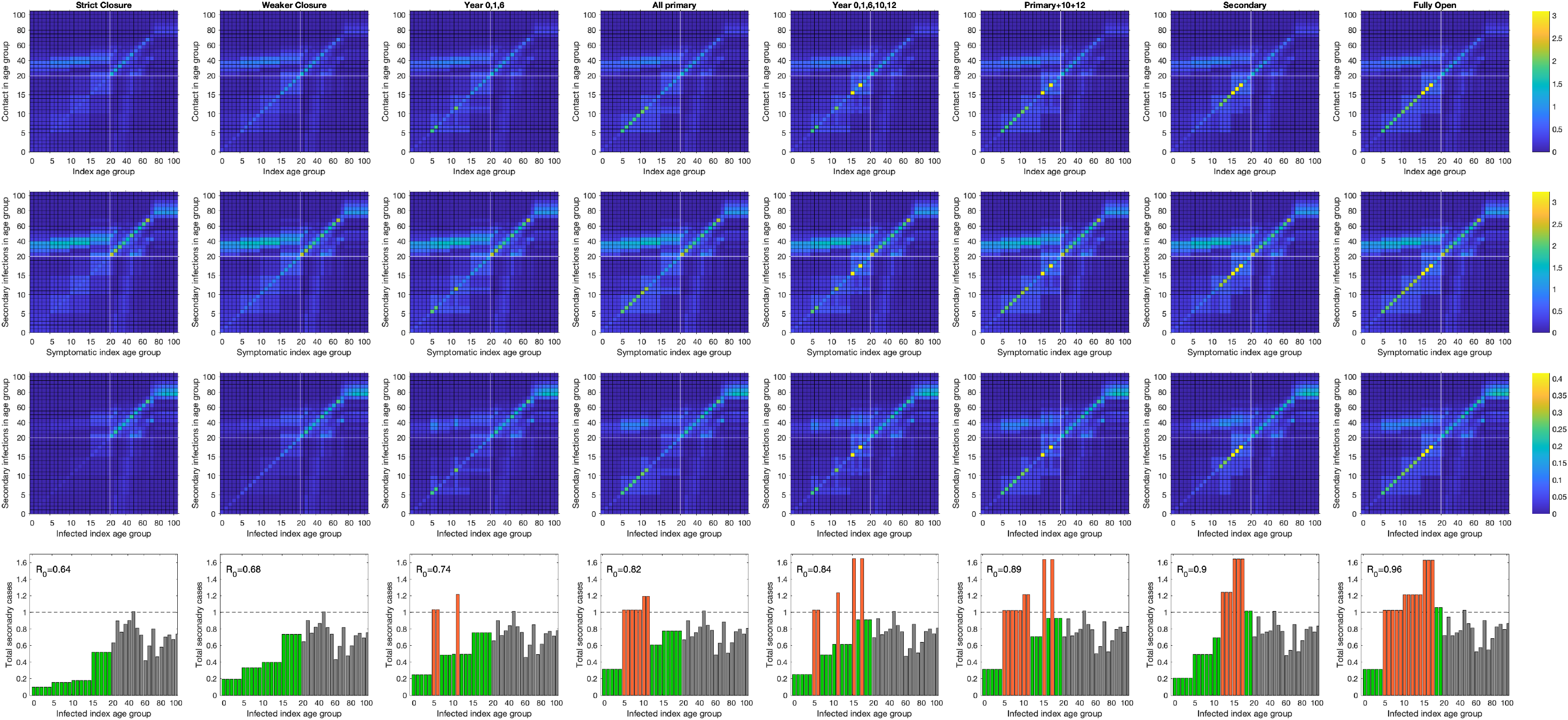
Mixing matrices and their implications for onwards transmission. We consider the effect on contact structures between different age groups under **(first column)** strict school closure, **(second column)** weak closure, **(third column)** years 0 (reception), 1 and 6 returning to school, **(fourth column)** all primary school children at school, **(fifth column)** years 0, 1, 6, 10 and 12 at school, **(sixth column)** all primary school children and years 10 and 12 at school, **(seventh column)** all secondary school children at school and **(eighth column)** all children in school. For each school closure we show: **(first row)** the average number of contacts by age for each index age group [36]; **(second row)** the average number of secondary infections for a symptomatic infected individual by age (combining the mixing matrix with age-susceptibility); and **(third row)** the average number of secondary infections for each infected individual by age combining the mixing matrix, age-susceptibility and the impact of asymptomatic transmission). **(Fourth row)** The total number of secondary infections for each infected index age group. Green bars indicate school year groups who remain at home, whilst red bars indicate year groups who return to school.

### Modelling school reopening scenarios

We used this model framework to evaluate eight strategies for reopening schools from 1st June. The eight school reopening options we considered assumed that, from the 1st June, the following school year groups would return to school:

i. reception (year 0), year 1 and year 6 (full class sizes);
ii. reception, year 1 and year 6 (half class sizes);
iii. all primary schools;
iv. reception, years 1, 6, 10 and 12 (full class sizes);
v. reception, years 1, 6, 10 and 12 (half class sizes);
vi. primary schools plus year groups 10 and 12;
vii. all secondary schools;
viii. all schools.

For clarity, in all the strategies considered here we assumed that children of key workers continued to attend school at the currently observed level.

We assessed the school reopening scenarios at a regional scale, modelling the population of England aggregated to seven regions (East of England, London, Midlands, North East and Yorkshire, North West England, South East England, South West England). This involved the use of region-specific posterior parameters obtained in our prior work, where we fit our base transmission model on a region-by-region basis, using a Monte Carlo Markov Chain (MCMC) fitting scheme, to four timeseries: (i) new hospitalisations; (ii) hospital bed occupancy; (iii) ICU bed occupancy; (iv) daily deaths (using data on the recorded date of death, wherever possible) [32]. The inference was performed from epidemiological data until 12th May 2020.

Our assessment of school reopening strategies comprised of three strands. Firstly, we quantified how the process of opening schools and year groups affected contact patterns and anticipated secondary infections. Secondly, we related the scale of school opening to the relative change in *R*, assuming the same transmission patterns in the rest of the population as during the strict lockdown phase. Finally, we gauged the estimated change in clinical case and its sensitivity to changes in community transmission following the easing of lockdown measures on 13th May. We outline each item in further detail below.

### Contacts and secondary infections

Any school reopening plan will inherently alter age-group contact patterns compared to contact structures observed during the lockdown. We attempted to resolve how these alterations in social interactions propagated into the transmission dynamics by tracking secondary infections arising from symptomatic index cases and infected index cases (either symptomatic or asymptomatic), respectively.

Specific to this aspect of the analysis we focused on a single region, namely the Midlands and the posterior parameter set with the maximum likelihood. We first assess the contact structure and transmission under two distinct lockdown assumptions (‘strict closure’ and our default assumption of ‘weaker closure’). The ‘strict closure’ scenario assumed that there was no additional mixing between school-age groups during the lockdown period. ‘Weaker closure’ assumed there was more limited adherence, leading to higher mixing between school-age groups compared to the ‘strict closure’ setting. We also consider six of the eight reopening strategies (omitting those with half class sizes as these are bounded above by the full-class strategy). For each we show the age-mixing matrix between age-groups; the transmission matrix from a symptomatic infectious individual; the transmission matrix from an average infectious individual (recognising the many will be asymptomatic or in household quarantine); and the expected number of secondary cases an average infectious individual of a particular age-group will generate.

### Reproduction number analysis

The reproduction ratio or number (*R*) has become a universally recognised quantity in the description of COVID-19 dynamics; it is defined as the average number of secondary cases from an average index case — where the second average is important as it samples across all infectious states including asymptomatics and those currently under household isolation. To prevent the occurrence of a second phase of exponential growth in infection, it is crucial that relaxation of social distancing measures do not result in the value of *R* rising above 1. On these grounds, there is interest in predicting the magnitude of a rise in *R* that could result from the reopening of schools, and our confidence in this result.

We considered all eight school reopening scenarios and examined the increase in *R* per region under each of the eight strategies. To compute *R*, we used the contact matrices associated with the given choice of school reopening and accounting for the regional population structure, whilst assuming the same level of mixing in the rest of the population as during the strict lockdown; therefore any changes in *R* are driven by changes in school-age mixing. We calculated means and intervals from 1000 simulation replicates with parameter sets sampled from the posterior parameter distributions.

### Clinical case impact

The prior methods focused on the reproduction number *R*, which is both an instantaneous measure (*R* can be calculated at any or every time point) and a long-term calculation (as it utilises an eigenvalue approach to generate the asymptotic *R*). Calculation of quantities of public health interest requires the simulation of the full temporal dynamics from the start of the outbreak to the closing of schools for the summer holidays on 22nd July. In addition, we considered the sensitivity of reopening schools to other potential changes in population mixing patterns (and hence different values of *R*) driven by other changes to the lockdown since 13th May. These changes to population mixing were generated by reducing the adherence with lockdown measures, bringing the mixing matrices closer to the prepandemic norm.

We performed these simulations, using the full dynamic model to generate estimates of the symptomatic cases, deaths and ICU admissions between 1st June and 22nd July, for each of the eight school-opening strategies. We compared these measures, aggregated over this 52-day period, to a scenario where school closures remain in place beyond the 1st June.

For each reopening strategy and each region, we performed a total of 1000 replicates. In each replicate we sampled parameter values randomly from all posterior parameter distributions, with the exception of the adherence level. The potential reduction in adherence values, from 13th May, inevitably generates different *R* values at the point of school reopening (measured by the observed growth rate of the outbreak in the model simulation). As a consequence, for comparative purposes we segregated the estimated increases in epidemiological quantities (comparing different school opening strategies for fixed underlying parameters) into three categories according to the *R* value before school reopening: below 0.8, between 0.8 and 1, or between 1 and 1.2.

## Results

### Choice of reopening strategy influences contact structure and secondary infection risk

We first investigated the impact of alternate strategies for reopening schools upon contact patterns between individuals and the effect of this upon transmission of SARS-CoV-2 and occurrences of COVID-19 infection. Our results for the Midlands, and the posterior parameter-set derived in May that maximises the likelihood (giving *R* ≈ 0.78), are summarised in Fig. 2. For all scenarios investigated we observe several common trends. Contacts are most common between individuals of the same, or similar ages (Fig. 2, first row [36]). There was also greater contact between children and adults between the ages of 25 and 55, reflecting interactions between children and their parents, as well as between elderly people [36]. This increased likelihood of contact within and between those age groups is reflected in the risk of secondary infections occurring (Fig. 2, second and third rows). The second row accounts for age-dependent susceptiblity, and shows the expected number of secondary infections in each age (y-axis) from a symptomatic index case of a particular age (x-axis). The third row incorporates the likely state of an index infection (symptomatic, asymptomatic or in household quarantine - as predicted by the underlying ODEs) thereby reducing the potential transmission from particular age-groups (Supporting Figure S2).

If schools remain closed, with a high level of adherence to the lockdown within this younger age-group (Fig. 2, first column) we observe that contact between children, and therefore the risk of secondary infection occurring, is extremely low. Should adherence to lockdown be weaker (Fig. 2, second column), we observe a higher rate of mixing between children and a slight increase in risk of secondary infections occurring. For both of these scenarios the average number of secondary infections per index infection is below 1 for all age groups and the value of *R* remains significantly below 1.

We now investigate the impact of various strategies for school reopenings. We first investigate the scenario of reception, year 1 and year 6 children returning to school - the policy that is scheduled to be implemented on 1st June in England (Fig. 2, third column). In this scenario, we observe a slight increase in contacts compared to the “weaker closure” scenario, with increased transmission between individuals in these age groups. However, crucially, even within these age groups, the total number of secondary infections per index case remains below one (third column, final row, red bars) and the overall reproduction number value of *R* was only observed to have slightly increased from the scenarios in which schools remain closed. A slight increase in mixing, and hence *R*, was again observed when all primary schools are opened (Fig. 2, fourth column), but we predict that *R* remains below 1.

To conclude this segment of the analysis, we investigated the impact of school reopening strategies that involved some, or all, secondary school children returning to the classroom. If children from key years of 10 and 12 return to school (in addition to some or all primary school children), a significant increase in mixing was observed within those age groups; the number of secondary infections as a result of index infections in secondary schools was predicted to be above one (Fig. 2, fifth and sixth columns). However, this expected number of cases is distributed across multiple age-groups thereby dissipating the worst effects. In general, we found secondary schools to represent a higher risk of increased transmission potential than primary schools. This could lead to higher values of *R* when all secondary schools are opened; but for all scenarios investigated, even the scenario in which all schools are opened, we found strong support for *R* remaining below 1 in the Midlands (Fig. 2, final column) assuming that all other transmission patterns remain unchanged.

### Effect of school reopening on reproduction number

Next, we sought to estimate changes in *R* that may result from school reopenings alone - assuming the transmission patterns in the rest of the population are maintained at strict lockdown phase levels. In contrast to the the first part of the analysis, which focused on a single set of parameters and a single region (Fig. 2), here we explore the full parameter uncertainty and compare different parts of the country.

For all school opening scenarios, within the seven regions of England, we observe an increase in *R* compared to what we predict for keeping schools closed until the end of the academic year (Fig. 3 and Supporting Figure S3). This is to be expected, given the increase in contact between children that such reopening scenarios would allow. However, the magnitude of increase is predicted to be relatively low, depending on the age-groups that return to school. In general, the more year groups allowed to return to school at one time, the greater the effect on *R*, with the return of secondary school children having the greatest impact.

**Fig. 3:**
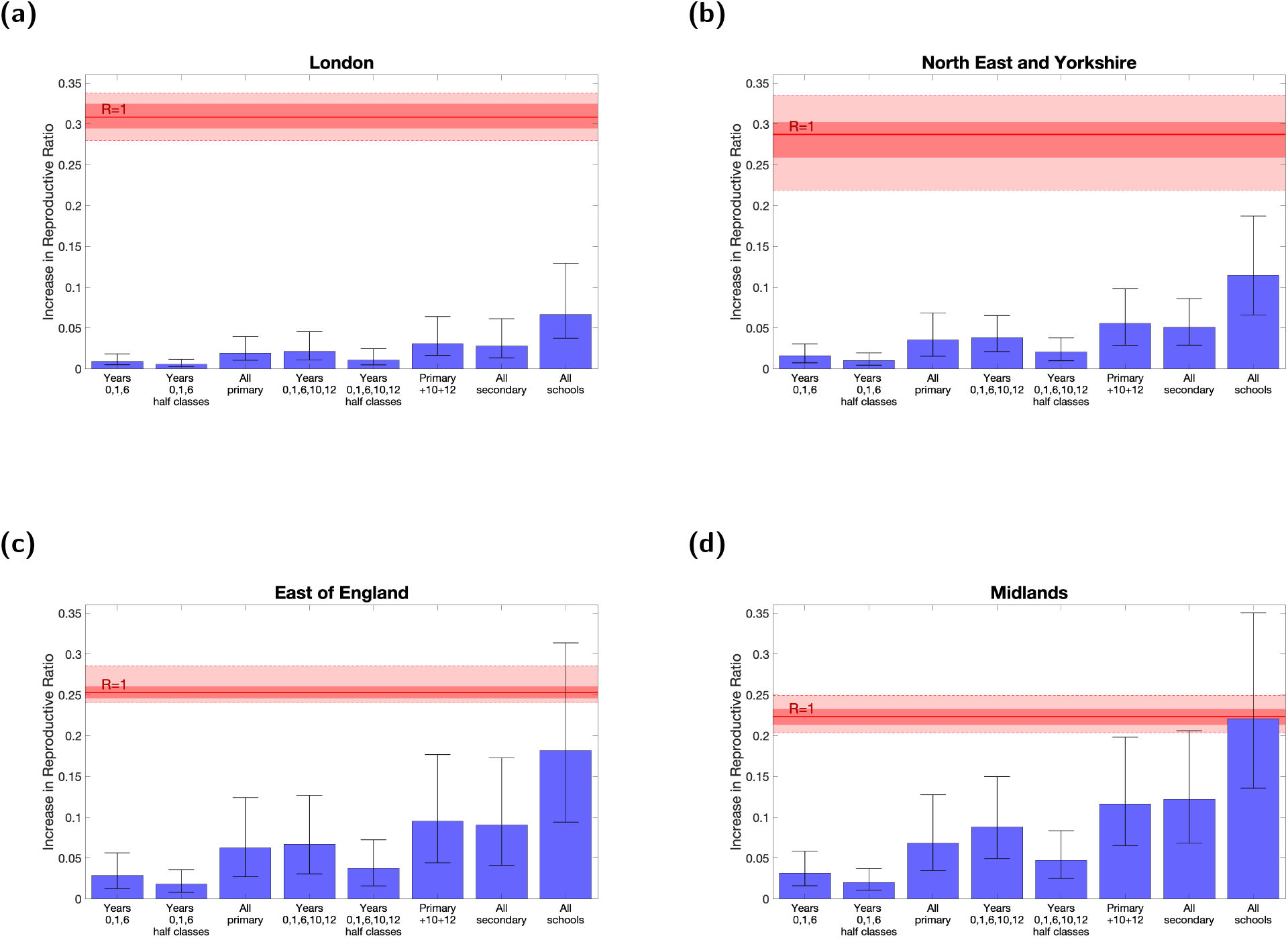
Increase in reproduction number, *R*, under eight school reopening scenarios for four regions in England. Estimates are depicted for the following four regions: **(a)** London (*R* ≈ 0.69), **(b)** North East and Yorkshire (*R* ≈ 0.71, **(c)** East of England (*R* ≈ 0.74), **(d)** the Midlands (*R* ≈ 0.78). For each scenario, bars represent the mean absolute increase in *R*, compared to what we would observe if schools remained closed. We also give the 95% prediction intervals. Solid red lines identify the absolute increase required to raise *R* above 1, within each region, alongside 50% and 95% intervals (shaded red areas). Means and intervals are calculated from 1000 replicates sampled from the posterior parameter distributions. All scenarios are implemented on 1st June and continued until 22nd July.

The impact of allowing multiple year groups to return to school can still be small: opening a fraction of the age-cohorts in each school generally leads to a moderate (less than 0.05) increase in *R*, especially if children can be taught in smaller class sizes which is assumed to lead to a proportionate reduction in within school transmission.

There is however considerable variation between the regions and here we focus on four exemplars. For London and North East England & Yorkshire, the increase in *R* was considerably less than that for East of England and the Midlands across all reopening scenarios. For the former, even allowing all age groups to return to school (while maintaining tight control in other age-groups) was highly unlikely to increase *R* above 1, with both means and 95% prediction intervals falling well below this threshold (Figs. 3(a) and 3(b), the 95% prediction intervals, as the name suggests, contain 95% of all predicted values across the entire posterior distribution of parameters). This low *R* value was especially true for London, which saw the most abrupt rise and subsequent decline in cases. However, this was not the case for the East of England (Fig. 3(c)) and the Midlands (Fig. 3(d)). In these regions, allowing schools to fully reopen could increase *R* above 1, with such an occurrence lying within the 95% prediction intervals. We attribute these regional differences to both heterogeneity in the observed rate of epidemic decline and the differential proportion of school age children in each region; the Midlands has the highest proportion of older teenagers in the country.

### Quantifying clinical case impact stemming from the re-opening of schools

Our final piece of analysis examined the extent to which each of the eight school reopening strategies may contribute to clinical case outcomes, using the full dynamic model. We also considered the sensitivity of reopening schools to other potential changes in population mixing patterns (and hence different values of *R*) driven by other changes to the lockdown since 13th May.

In each scenario, reopening schools increased the absolute number of cases, ICU admissions and deaths as a result of increased transmission (Fig. 4). Note that these increases will not be restricted to the children that return to school, since the greater transmission will lead to increased cases in other age groups. Echoing our earlier findings, strategies in which a larger number of children return to school generally resulted in larger increases. In addition, older children had a greater effect, so that reopening secondary schools results in larger increases than only reopening primary schools.

**Fig. 4:**
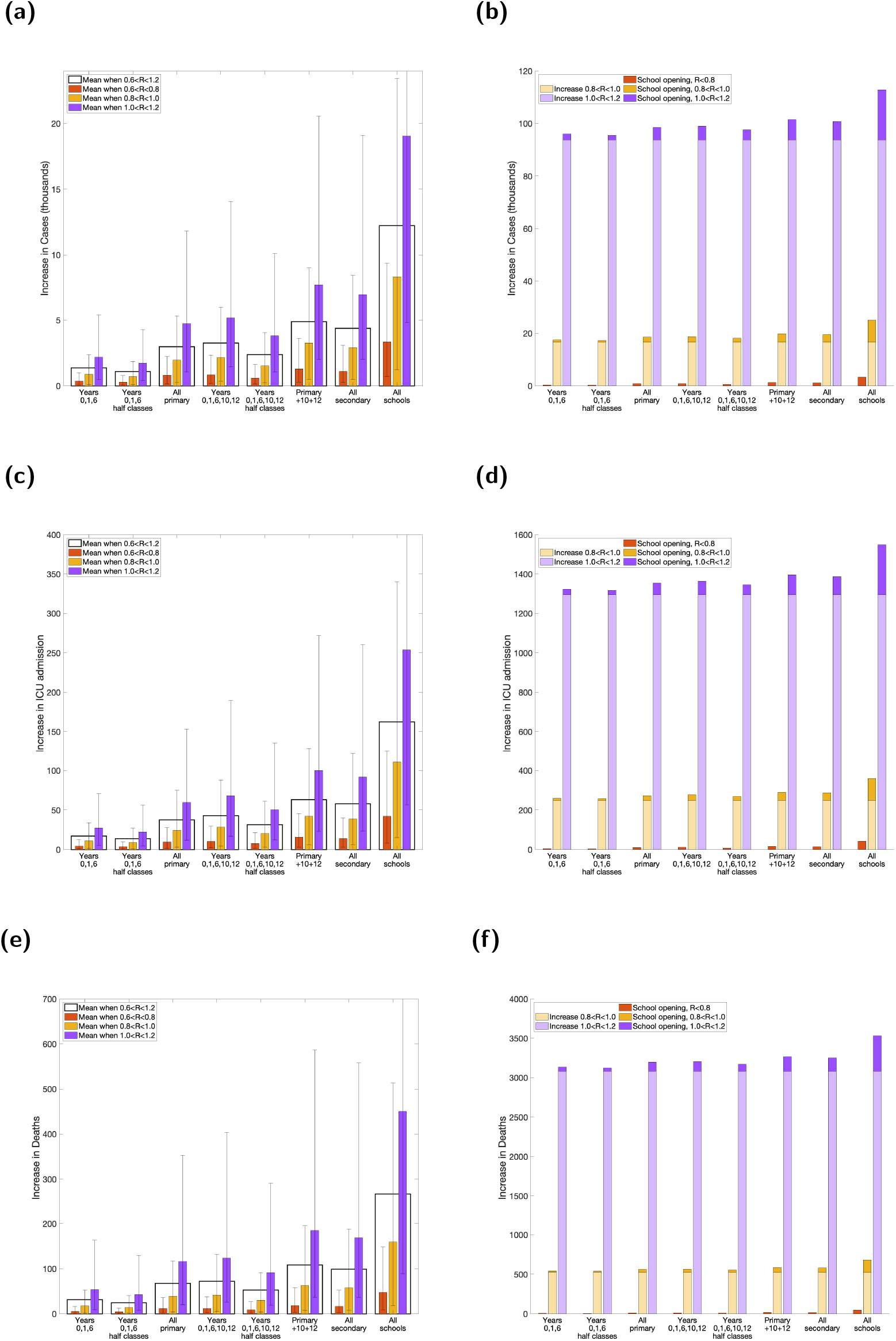
Increase in disease burden and clinical case outcomes from 1st June to 22nd July under the eight different scenarios representing various combinations of school years return to school. **(a,b)** Cases; **(c,d)** ICU admissions and **(e,f)** deaths. For each scenario, the three coloured bars give the increase relative to if no schools returned for low (red), intermediate (yellow) and high (purple) reproduction numbers, while the clear bar (in panels a, c and e) is the mean across all reproduction numbers. Prediction intervals are given for each scenario representing the uncertainty in the predicted values. In panels b, d and f, we also display (in lighter colours) the increase in each quantity that is associated with the change in *R* from the current low situation.

The opening of schools on 1st June was just one of a collection of changes in a short space of time, which began from 13th May. In the previous sections we focused on school reopening, assuming that mixing (and hence transmission) within the wider population remained unchanged. Here we allowed the relaxation of lockdown measures to precipitate an increase of *R* within the community and calculate the additional change from the opening of schools. We consistently found that school reopening had a larger impact when *R* in the community was high, leading to a greater increase in cases, ICU admissions and deaths. However, by far the largest increase in any of these key quantities was driven by the underlying change in *R* due to relaxations other than the reopening of schools (Figs. 4(b), 4(d) and 4(f)).

## Discussion

In this paper, we have described a mathematical model used in May 2020 to consider the implications of various potential strategies for reopening schools in England. We have compared the different strategies by presenting mixing matrices and discussing their implication for onward transmission, and by analysing the increase in the reproduction number and absolute number of cases, ICU admissions and deaths compared to those predicted if schools remain closed. Given that in May all regions were estimated to have reproduction numbers (*R*) below 0.8, we predicted that, in the absence of other changes, the complete opening of all schools was unlikely to raise the reproduction number above one. It must be noted that even though *R* remains below one, the slight increase in transmission resulting from school reopening subsequently leads to a small increase in the absolute number of cases, ICU admissions and deaths. If the reopening of schools is part of a wider policy of relaxing controls, then the impact of these additional changes must also be factored into the analysis [30, 31].

Reopening schools, in any form, inevitably leads to more mixing between children, an increase in *R* and thus more transmission of the disease. However, we can constrain and potentially minimise the extent of this increase by selecting a subset of year-groups to return to school. In doing so, we restrict the increase in *R* to very low levels and, crucially, avoid any possibility of increasing *R* above 1. These findings are in agreement with studies applied to other nations suggesting that school settings are not a major driver of SARS-CoV-2 transmission. A statistical study in US counties looking at the relationship between the reduction in growth rate and the timing of different state and local government social distancing interventions found school closures to not be statistically significant [37]. Further, in terms of suppressing spread of SARS-CoV-2, a mechanistic transmission model evaluating the impact of non-pharmaceutical interventions in Switzerland, by their potential to reduce *R* below 1 at a national level, predicted school closures alone would typically be insufficient to induce control [38].

In choosing a specific reopening policy, decision-makers must weigh-up the benefits to both children and parents that are gained from allowing more year groups to return to school, with the risks associated with increased transmission. In light of the variation in effects on *R* between regions, reopening policies may benefit from heterogeneity across the country, in order to allow the most children possible to return to school without threatening a resurgence of disease prevalence. Our results also highlighted the benefit to be gained from small class sizes and hence maintaining such measures of social distancing, with the impact of this form of non-pharmaceutical intervention within the school environment difficult to infer without explicit data.

Our results also predicted a higher risk of increased transmission associated with reopening secondary schools compared to the reopening of primary schools. Such a relationship may be partly attributed to the observed larger number of contacts of secondary school children compared to primary school children [36]. Additionally, other contributory factors include differences between age groups in terms of susceptibility and, if infected, displaying symptoms [7, 8]. These may consequently lead to secondary school children having a larger contribution to overall transmission throughout the population. This could potentially be offset by the greater ability of older pupils to understand and abide by social distancing advice.

Increasing levels of contacts between school children inevitably leads to greater absolute numbers of infections, detected cases, ICU admissions and, regrettably, deaths, even if the reproduction number is not raised above one. For this reason, we also estimate the increase in these outcomes as a result of reopening schools using the different strategies. The ranking of the different strategies for these outcomes mirrors the ranking in terms of increases in *R*. The epidemiological impact of reopening schools also depends on the behaviour of the wider population. If there is more mixing within the adult (and elderly) population, the effect of reopening schools will be exacerbated by the generally higher infection levels and contacts in the community. Reopening schools will then lead to greater increases in case numbers over and above the increases due to greater mixing. In general, we found that even small changes in *R* due to the behaviour of the general population swamp the impacts of reopening schools. We would stress that, such increases must be viewed in the context of the restrictions currently placed on pupils and parents. Ultimately, it is a societal decision to balance the benefits to pupils’ welfare and education against the epidemiological consequences.

To consider the effects of specific school years returning, this work made some simplifying assumptions, and our results therefore have limitations. In particular, in this paper we consider only an England-specific context. The devolved administrations employ a different school system from England, including different school term dates, which may affect the outcome of reopening schools. Future work could incorporate such differences, some of the epidemic variability between nations will be captured by the model parameter fits that are already performed for all the devolved nations. In our analysis of schools we have made the pessimistic assumption that there will be limited non-pharmaceutical intervention within the school setting, however, we have ignored the potentially greater mixing of parents or other adults when taking younger children to school. Also, the model is deterministic, and captures the return to school in terms of increased mixing between school ages; it cannot capture the inevitable heterogeneity between schools, with some schools experiencing many cases while others have none. Similarly, we make no attempt to replicate the reactive closure of classes to prevent further spread once cases are identified.

As we have shown, the context in which school reopening happens will also have an impact on its effect. While we consider different population level mixing patterns, this exploration is necessarily constrained; for example it may also be the case that the opening of schools allows more parents to return to work, increasing their risk of infection [30]. Indeed, a surge in cases in Seoul, South Korea linked to a distribution centre, has identified at least one SARS-CoV-2 positive high-school student, whose family member worked at the centre; this was followed by the re-implementation of localised lockdown and social distancing measures, including the closure of 251 schools, days after their phased reopening [39]. It is also be important to consider the impact of school re-openings in combination with other concurrent measures, such as the NHS test and trace system in England (that began on 28th May) [40], which aims to trace close recent contacts of anyone who tests positive for SARS-CoV-2 and, if necessary, notify them to self-isolate at home to prevent onward transmission. Effective contact tracing breaks transmission chains, but may also subject school classes to tracing and isolation. Even without national-scale relaxation in the lockdown measures, the behaviour of the general population is likely to change over time, in ways that are difficult to predict. Beyond these considerations, we have also neglected the many possible side effects of reopening schools, such as parents interacting at the school gates, teachers’ exposure while travelling to school (or in the staff room), or the effects of school reopening on children mixing outside of school.

These analyses, performed in May, indicated that it should have been feasible to reopen all schools in June. Reopening schools (for June and July) while other measures remained constant would have allowed accurate information regarding the impact of children returning to the classroom for a relatively short period, and would have provided invaluable evidence on the role of younger age-groups in transmission. In practice, there was only a partial reopening on 1st June, with reception (year 0), year 1 and year 6 returning to primary schools, and the sporadic return of some years 10 and 12 to senior schools from 15th June. We predicted that the general return of just three primary school years would have a minimal impact on *R*, and very few school-based outbreaks were reported before the main summer holidays [41]. Unfortunately, before the return of all children to school in September, multiple regions (notably Leicester, Manchester and the North West) of England experienced high case numbers, while *R* continued to rise - such that it was likely to be above 1 before the reopening of schools. However, our modelling still generates some useful predictions. School reopening is predicted to have a larger impact in September than it would have done in June, although the impact is still relatively small compared to other relaxations of lockdown. Our work also suggests that measures to mitigate the rise in cases as we approach winter would be best focused on other routes of transmission, as even when *R* is significantly above 1, the effect of opening or closing schools is minimal.

## Data Availability

Data on cases were obtained from the COVID-19 Hospitalisation in England Surveillance System (CHESS) data set that collects detailed data on patients infected with COVID-19. Data on COVID-19 deaths were obtained from Public Health England. These data contain confidential information, with public data deposition non-permissible for socioeconomic reasons. The CHESS data resides with the National Health Service (www.nhs.gov.uk) whilst the death data are available from Public Health England (www.phe.gov.uk).

## Acknowledgements

The authors would like to thank Massimiliano Tamborrino for contributing to data curation, and the RAMP Rapid Review Group for their useful comments on this manuscript.

## Author contributions

**Conceptualisation:** Matt J. Keeling.

**Data curation:** Matt J. Keeling; Glen Guyver-Fletcher; Alexander Holmes.

**Formal analysis:** Matt J. Keeling.

**Investigation:** Matt J. Keeling.

**Methodology:** Matt J. Keeling.

**Software:** Matt J. Keeling; Edward M. Hill; Louise Dyson; Michael J. Tildesley.

**Validation:** Matt J. Keeling; Edward M. Hill; Benjamin D. Atkins; Louise Dyson; Michael J. Tildesley.

**Visualisation:** Matt J. Keeling.

**Writing - original draft:** Michael J. Tildesley; Edward M. Hill; Louise Dyson; Benjamin D. Atkins; Matt J. Keeling; Bridget Penman; Erin Gorsich; Emma Southall.

**Writing - review & editing:** Matt J. Keeling; Edward M. Hill; Louise Dyson; Benjamin D. Atkins; Erin E. Gorsich; Bridget Penman; Glen Guyver-Fletcher; Alexander Holmes; Hector McKimm; Emma Southall; Michael J. Tildesley.

## Ethical considerations

The data were supplied from the CHESS database after anonymisation under strict data protection protocols agreed between the University of Warwick and Public Health England. The ethics of the use of these data for these purposes was agreed by Public Health England with the Government’s SPI-M(O) / SAGE committees.

## Supporting information items

**Supporting Text S1**

Description of the complete system of model equations.

**Supporting Text S2**

Details on the mechanisms underpinning social distancing measures within the model framework

**Supporting Figure S1**

**Posterior distributions of key model parameters from fitting to date until 1st June**. The left-hand graphs show how the probability of symptoms (*d_a_*) and susceptibility (*σ_a_*) varies with age; given the low value of *alpha* most of the age-dependence is in the displaying of symptoms. The right-hand graph shows the relative adherence with lockdown measures in each region; high values correspond to a dramatic reduction in the mixing matrix, while an adherence of zero returns the matrix to pre-lockdown levels. This figure supplements the information in Table 1. Bars show the 95% credible intervals from the posterior distribution.

**Supporting Figure S2**

**Distribution of household symptomatic, asymptomatic and isolated cases in each age group on 1st June**. Used in conjunction with Fig. 1. Bottom segments (blue shading) represent symptomatic infection. Middle segments (orange shading) represent asymptomatic infection. Top segments (yellow shading) represent those in isolation. Filled dots specify the fraction of the population within that age bracket.

**Supporting Figure S3**

**Increase in reproduction number, *R*, under eight school reopening scenarios for three regions in England**. Estimates are depicted for the following three regions: (**a**) North West, (**b**) South East, (**c**) East of England. For each scenario, bars represent the mean absolute increase in R, compared to what we would observe if schools remained closed. We also give the 95% prediction intervals. Solid red lines identify the absolute increase required to raise *R* above 1, within each region, alongside 95% credible intervals (dashed red lines). Means and intervals are calculated from 1000 replicates sampled from the posterior parameter distributions. All scenarios are implemented on 1st June and continued until 22nd July.

